# Synthetic Magnetic Resonance Spectra Generation using convex NMF in Neuro-Oncology

**DOI:** 10.64898/2026.11.24.26368829

**Authors:** Maitiú Mac Cárthaigh, Lili F. Tóth, Alfredo Vellido, Margarida Julià-Sapé

**Affiliations:** Institut de Biotecnologia i Biomedicina, Universitat Autònoma de Barcelona, Cerdanyola del Vallès, Spain; Departament de Bioquímica i Biologia Molecular, Universitat Autònoma de Barcelona, Cerdanyola del Vallès, Spain; CIBER de Bioingeniería, Biomateriales y Nanomedicina, CIBER-BBN, Spain; Department of Computer Science, Universitat Politècnica de Catalunya (UPC), Barcelona, Spain; Intelligent Data Science and Artificial Intelligence Research Center, IDEAI-UPC, Barcelona, Spain

**Keywords:** magnetic resonance spectroscopy (MRS), synthetic data generation, convex non-negative matrix factorisation (cNMF), brain tumour classification, Gaussian mixture models (GMM), kernel density estimation (KDE), linear discriminant analysis (LDA)

## Abstract

Magnetic Resonance Spectroscopy is a powerful, non-invasive technique for investigating the metabolic composition of biological tissues, particularly useful in the classification of brain tumours. However, its inclusion into machine learning pipelines is limited by the scarcity of large, labelled datasets. This work evaluates whether synthetic MR spectra generated using convex Non-negative Matrix Factorisation and probabilistic modelling of the resulting weight distributions can improve brain tumour classification. MR spectra from the INTERPRET database were decomposed into interpretable source and weight matrices, both separately for each of six diagnostic tasks and jointly across five tissue classes, at short and long echo time. Class-specific weight distributions were modelled using Gaussian Mixture Models and Kernel Density Estimation across a range of hyperparameter settings, and new weight vectors were sampled and recombined with the fixed source matrix to produce synthetic spectra; hyperparameters were selected by the mean squared error to the real class means. The diagnostic utility of the synthetic spectra was assessed with Linear Discriminant Analysis under three internal and four external evaluation strategies, the latter using an independent multi-centre cohort. Synthetic spectra were readily classified by models trained on real data (mean accuracy 0.87), but models trained on synthetic spectra alone generalised poorly, both to real INTERPRET spectra (0.55) and to the external cohort (0.47, against 0.84 for training on real data; ***p ≤* 0.003**). Augmenting real training data with synthetic spectra neither improved nor significantly harmed external accuracy (mean paired differences between ***−*0.04** and **+0.01**; all ***p ≥* 0.027** uncorrected, none significant after correction), and class-balanced augmentation did not help, and task-specific and joint decompositions performed comparably. The selected generative models were low in complexity and produced spectra closer to the class means than the real data are, suggesting that selection by reconstruction fidelity is poorly aligned with the goal of augmentation, and that internal validation alone would have overstated the value of the synthetic data.

## 1 Introduction

### Clinical context and motivation

Primary brain and other central nervous system (CNS) tumours are, in absolute terms, an uncommon disease group. Across the 40 countries covered by the most recent European Cancer Information System estimates, brain and CNS tumours accounted for an estimated 67,100 new diagnoses in 2020, corresponding to an age-standardised incidence rate of 8.9 per 100,000 population; within the EU-27 specifically, the corresponding figures were 43,600 new cases and an age-standardised rate of 9.4 per 100,000 [1]. This means that brain and CNS tumours are far from the most common types of cancer, yet their impact is disproportionate to their rate of incidence; histological diversity is high, prognosis varies greatly between tumour types and grades, and treatment decisions depend on accurate, early characterisation of the mass.

Tissue biopsy remains the diagnostic gold standard but is invasive, carries surgical risk, and is not always feasible depending on tumour location - especially true in the case of brain tumours. This has motivated interest in non-invasive diagnostic techniques capable of discriminating tumour type without surgical intervention, and proton magnetic resonance spectroscopy (^1^H-MRS) is one such technique, providing a biochemical fingerprint of tissue metabolism that complements conventional structural imaging. Metabolite patterns derived from MRS have been repeatedly linked to tumour type and grade. Reliable automatic classification from MRS data therefore has direct clinical value, potentially shortening the diagnostic pathway and reducing reliance on invasive procedures.

### The data scarcity problem in MRS

Realising the clinical potential of machine learning for MRS is limited by the availability of large and well-labelled training data. Compared with structural Magnetic resonance Imaging (MRI), MRS acquisition is technically demanding: spectra are more sensitive to field inhomogeneity, require careful voxel placement and longer acquisition times. They are also more susceptible to motion, baseline artefacts, and low signal-to-noise ratios. As a result, MRS is acquired far less routinely in clinical practice and is often reserved for cases where conventional imaging is inconclusive.

The spectra that are acquired must also undergo expert quality control before they can be used for model development [2]. Poor shimming, lipid contamination, and other acquisition artefacts frequently leave us with spectra unsuitable for analysis, further reducing the number of usable samples. In addition, while brain tumours at large are uncommon, many diagnostic subtypes are rarer still. Building datasets of sufficient size therefore requires pooling data from multiple clinical centres, introducing additional variability due to differences in scanners, acquisition protocols, and patient populations.

Consequently, even the largest collaborative MRS datasets contain few high-quality spectra per diagnostic class. The INTERPRET [3] and eTumour [4] datasets used here (Sections 2.1.1, 2.1.2) provide only a few dozen samples per class once quality control and echo time are accounted for. Models trained on datasets of this size overfit and often fail to generalise to unseen centres; an early evaluation of INTERPRET-trained classifiers on eTumour spectra was the first of its kind, motivated by the absence of cross-centre validation in the field [5]. Improving cross-centre generalisation, rather than maximising internal accuracy, is therefore the key challenge and motivates the external validation used here.

### Synthetic Spectral Augmentation: Promise and Open Questions

Synthetic data generation is one of several strategies that have been proposed to address data scarcity in settings where acquiring further real, labelled examples is impractical or prohibitively expensive. Rather than acquiring more spectra, the aim is to model the underlying distribution of existing real data closely enough that additional, plausible samples can be drawn from it and used to augment a limited training set, either to increase effective sample size, to rebalance under-represented classes, or both. Applied naively, this strategy carries an obvious risk: if the generative model captures the real data distribution poorly, or captures it too narrowly, synthetic samples may fail to add genuine variability, may introduce artefacts of their own, or may simply reproduce the biases already present in the limited real training set, in which case augmentation would be expected to help internal validation metrics without conferring any real benefit - or even causing harm - when the resulting classifier is evaluated on genuinely independent data.

Whether or not synthetic augmentation boosts external generalisation, as opposed to merely increasing internal cross-validation performance, is therefore an empirical question that must be evaluated directly against held-out data acquired under different conditions from the training set. A small number of prior studies have explored generative approaches specifically for MRS data of brain tumours [6–12], though, to our knowledge, none has evaluated a convex-Non-negative Matrix Factorisation (cNMF)-based generation strategy of the sort proposed within this study – the closest methodological precedent, cNMF-based decomposition of MRS data for source extraction and dimensionality-reduction-driven classification [13–15], uses the fitted decomposition directly for classification rather than to generate new synthetic training samples – nor, to our knowledge, benchmarked synthetic augmentation against a fully independent, multi-centre external cohort of the kind used in this study.

### Purpose of the study

The purpose of this study is to determine whether synthetic MRS spectra, generated via cNMF decomposition and weight-space resampling, can improve the external generalisation of brain tumour classifiers beyond what is achievable using internal real training data alone. Specifically, we (i) decompose real INTERPRET spectra into class-specific source and weight matrices via convex NMF, both independently per experiment and jointly across all five diagnostic classes; (ii) model the resulting weight distributions using Gaussian Mixture Models (GMM) and Kernel Density Estimation (KDE), generating synthetic spectra across a range of hyperparameter settings and selecting the best-performing configuration by reconstruction fidelity; (iii) evaluate the internal validity of the resulting synthetic data across three complementary classification strategies, assessing whether synthetic spectra maintain compatible distributions with real data and whether they can substitute for or augment it; and (iv) assess, across six diagnostic experiments and two echo times, whether augmenting real training data with synthetic spectra improves classification performance on a fully independent, multi-centre external test cohort relative to a real-data-only baseline. In doing so, this study aims to provide direct evidence on whether cNMF-based synthetic augmentation is a viable route to improving the clinical generalisability of MRS-based brain tumour classifiers, rather than a strategy that merely improves apparent performance under internal validation.

## 2 Methods

### 2.1 Datasets

#### 2.1.1 INTERPRET Dataset

The primary dataset used for model training and internal validation was sourced from the INTERPRET project (2000–2002) [3], a multi-centre collaboration that acquired ^1^H-MRS and associated clinical data from brain tumour patients. The dataset originates from a subset of cases that passed all quality controls and consists of 304 patients with single-voxel (SV) spectra acquired under standardised protocols across multiple institutions, at short echo time (STE, TE=20–32 ms) and/or long echo time (LTE, TE=135–144 ms), and has been used in multiple studies [5, 16–21]. The dataset encompasses five diagnostic categories relevant to this study: astrocytoma grade II (A2), glioblastoma (GL), metastasis (ME), meningioma (MM), and normal brain tissue (NO). Class sizes ranged from 15 to 86 spectra per class per echo time, reflecting the inherent scarcity of labelled clinical MRS data. A full breakdown of sample counts per class and echo time is provided in Table 1. Table 2 summarises the acquisition parameters according to the MRSinMRS reporting standards [22].

**Table 1.** Number of spectra available per diagnostic class and echo time for the INTERPRET and eTumour datasets.

|  | **INTERPRET**<br>(Training set) |  | **eTumour**<br>**IDI-Bellvitge**<br>(Test set) |  |
|---|---|---|---|---|
| **Class** | **STE** | **LTE** | **STE** | **LTE** |
| Astrocytoma grade II (A2) | 22 | 20 | 10 | 10 |
| Glioblastoma (GL) | 86 | 78 | 23 | 23 |
| Metastasis (ME) | 38 | 31 | 9 | 9 |
| Meningioma (MM) | 58 | 55 | 10 | 10 |
| Normal (NO) | 22 | 15 | 5 | 5 |
| **Total** | **226** | **199** | **57** | **57** |

**Table 2.**
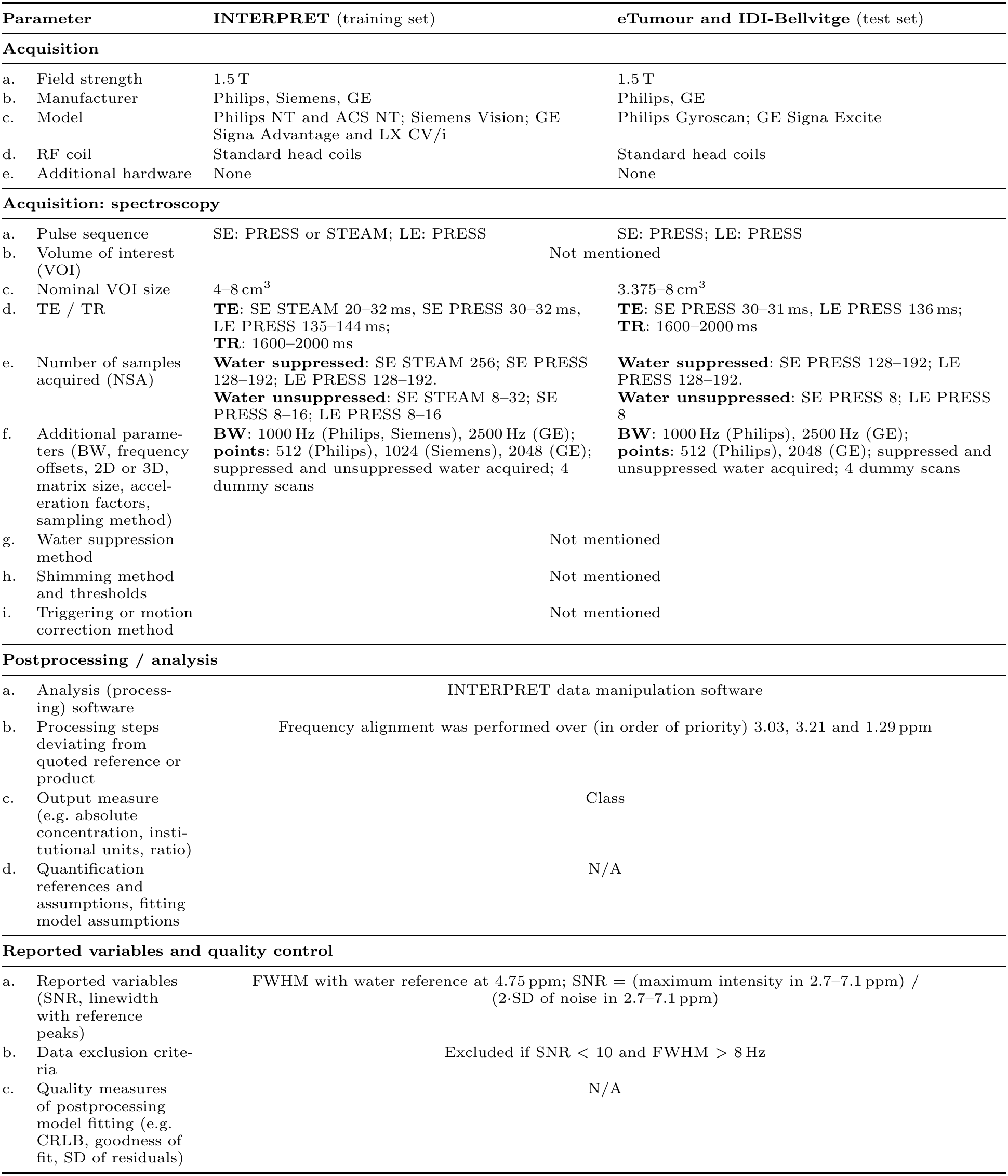
Acquisition parameters of the training (INTERPRET) and test (eTumour, IDI-Bellvitge) datasets, reported according to the recommended MRSinMRS standards [22]. SE: short echo; LE: long echo.

| Parameter |  | **INTERPRET** (training set) | **eTumour and IDI-Bellvitge** (test set) |
|---|---|---|---|
| **Acquisition** |  |  |  |
| a. | Field strength | 1.5 T | 1.5 T |
| b. | Manufacturer | Philips, Siemens, GE | Philips, GE |
| c. | Model | Philips NT and ACS NT; Siemens Vision; GE Signa Advantage and LX CV/i | Philips Gyroscan; GE Signa Excite |
| d. | RF coil | Standard head coils | Standard head coils |
| e. | Additional hardware | None | None |
| **Acquisition: spectroscopy** |  |  |  |
| a. | Pulse sequence | SE: PRESS or STEAM; LE: PRESS | SE: PRESS; LE: PRESS |
| b. | Volume of interest (VOI) | Not mentioned |  |
| c. | Nominal VOI size | 4–8 cm<sup>3</sup> | 3.375–8 cm<sup>3</sup> |
| d. | TE / TR | **TE**: SE STEAM 20–32 ms, SE PRESS 30–32 ms, LE PRESS 135–144 ms;<br>**TR**: 1600–2000 ms | **TE**: SE PRESS 30–31 ms, LE PRESS 136 ms;<br>**TR**: 1600–2000 ms |
| e. | Number of samples acquired (NSA) | **Water suppressed**: SE STEAM 256; SE PRESS 128–192; LE PRESS 128–192.<br>**Water unsuppressed**: SE STEAM 8–32; SE PRESS 8–16; LE PRESS 8–16 | **Water suppressed**: SE PRESS 128–192; LE PRESS 128–192.<br>**Water unsuppressed**: SE PRESS 8; LE PRESS 8 |
| f. | Additional parame-ters (BW, frequency offsets, 2D or 3D, matrix size, accel-eration factors, sampling method) | **BW**: 1000 Hz (Philips, Siemens), 2500 Hz (GE); **points**: 512 (Philips), 1024 (Siemens), 2048 (GE); suppressed and unsuppressed water acquired; 4 dummy scans | **BW**: 1000 Hz (Philips), 2500 Hz (GE); **points**: 512 (Philips), 2048 (GE); suppressed and unsuppressed water acquired; 4 dummy scans |
| g. | Water suppression method | Not mentioned |  |
| h. | Shimming method and thresholds | Not mentioned |  |
| i. | Triggering or motion correction method | Not mentioned |  |
| **Postprocessing / analysis** |  |  |  |
| a. | Analysis (process-ing) software | INTERPRET data manipulation software |  |
| b. | Processing steps deviating from quoted reference or product | Frequency alignment was performed over (in order of priority) 3.03, 3.21 and 1.29 ppm |  |
| c. | Output measure (e.g. absolute concentration, insti-tutional units, ratio) | Class |  |
| d. | Quantification references and assumptions, fitting model assumptions | N/A |  |
| **Reported variables and quality control** |  |  |  |
| a. | Reported variables (SNR, linewidth with reference peaks) | FWHM with water reference at 4.75 ppm; SNR = (maximum intensity in 2.7–7.1 ppm) / (2·SD of noise in 2.7–7.1 ppm) |  |
| b. | Data exclusion crite-ria | Excluded if SNR < 10 and FWHM > 8 Hz |  |
| c. | Quality measures of postprocessing model fitting (e.g. CRLB, goodness of fit, SD of residuals) | N/A |  |

#### 2.1.2 External Test Set

To assess the generalisation of classifiers trained on synthetic data to an entirely unseen cohort, a complementary multi-centre SV MRS tumour dataset, acquired under comparable protocols at STE and LTE was used. Tumour spectra were acquired as part of the EU-funded eTumour project[4, 14, 23], at three clinical centres in the Barcelona metropolitan area: CETIR-CDP (Centre Diagnòstic Pedralbes, Unitat Esplugues, Esplugues del Llobregat, Spain), CRC-Corporacío Sanitària–IAT (Institut d’Alta Tecnologia, Barcelona, Spain), and IDI-Badalona (Institut de Diagnòstic per la Imatge, Unitat Badalona, Badalona, Spain). These data had previously been used as an independent test set in two studies[17, 23] Additionally, the normal brain test data had been acquired at Institut de Diagnòstic per la Imatge, Unitat Bellvitge, L’Hospitalet de Llobregat, Spain, and were also previously reported [24]. None of the test set spectra were used at any stage of model training, feature selection, or hyperparameter optimisation, ensuring a fully independent evaluation. The same preprocessing and normalisation procedures applied to the INTERPRET data were applied identically to the external spectra prior to evaluation. Class sizes for the external test set after label harmonisation are provided in Table 1. Acquisition parameters are also detailed in Table 2.

#### 2.1.3 Preprocessing and Spectral Representation

Raw data had been previously processed according to the INTERPRET protocol [23, 25, 26], which, in brief, includes: Fourier transform; division of the suppressed water by the unsuppressed water spectrum; line broadening (1Hz) water suppression with the HLSVD algorithm (10 components in the x y interval); residual water removal setting spectral points between 4.2 and 5.1 ppm to zero; linear interpolation to 512 points over a 1000Hz of BW, resulting in 512-point vectors spanning *−*2.7 to 7.1 ppm, followed by L2-normalisation. For this work, spectra were further cropped to a fixed-length vector of 235 frequency bins subsequently L2-normalised, such that the Euclidean norm of each spectrum equalled one [27]. This normalisation was applied independently to each spectrum and consistently across real, reconstructed, and synthetic samples.

#### 2.1.4 Experimental Configurations

Because the value of synthetic augmentation is likely to depend on the difficulty and structure of the classification task, we did not evaluate it on a single problem but across six binary and multi-class tasks spanning a range of clinical difficulty, from a high-separability baseline to overlapping tumour-only comparisons:

- **A2 vs NO** — binary discrimination between low-grade astrocytoma and normal brain tissue, serving as a high-separability baseline.
- **MM vs ME** — binary classification between meningioma and metastasis, representing a more challenging tumour-only comparison with overlapping metabolic profiles.
- **MM vs ME vs A2** — three-class tumour-only classification combining two spectrally distinct low-grade tumour types, meningioma and low-grade astrocytoma and metastasis, which is high grade.
- **MM vs ME vs NO** — three-class task introducing healthy tissue alongside two tumour types, one low grade and the other high grade.
- **MM vs GL vs A2** — three-class tumour-only classification combining two spectrally distinct low-grade tumour types, meningioma and low-grade astrocytoma and glioblastoma, a high-grade tumour with a population-diverse pattern.
- **MM vs GL vs NO** — three-class experiment combining low-grade beingn tumour, high-grade class-diverse tumour, and normal brain tissue.

Each configuration was evaluated independently for both STE and LTE acquisitions, yielding twelve distinct classification settings in total. This design allows us to test not only whether augmentation helps on average, but whether any benefit is consistent across task difficulty and echo time, or confined to particular regimes.

### 2.2 Representing Class Structure via cNMF

The first requirement for generating synthetic spectra is a representation in which each class’s variability can be modelled directly. A 235-dimensional spectrum is too high-dimensional to model class-conditionally from only a few dozen examples; we therefore seek a low-dimensional representation that (i) captures the dominant spectral variation within each class, (ii) remains physically interpretable as MRS signal, and (iii) permits exact reconstruction back to spectrum space so that samples drawn in the low-dimensional space can be turned back into spectra. cNMF [28] satisfies all three requirements, which motivates its use here.

#### 2.2.1 Convex NMF

cNMF was applied independently to each experimental subset to decompose the spectral data matrix **V** *∈* R*^m×n^*, where *m* = 235 denotes the number of frequency bins and *n* the number of spectra, into a product of source and weight matrices:

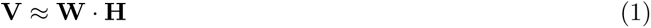

where **W** *∈* R*^m×k^* contains *k* basis spectra and **H** *∈* R*^k×n^* encodes the per-sample contribution of each source component. Unlike standard NMF, cNMF constrains **W** to lie within the convex hull of the original data, such that **W** = **VA** for some non-negative matrix **A** *≥* 0 with columns summing to one [13, 29]. This constraint encourages basis vectors to resemble convex combinations of real spectral profiles rather than abstract mathematical constructs, securing our requirement (ii) above, and improving the interpretability of the decomposition and ensuring that extracted sources remain physically plausible as MRS spectra.

#### 2.2.2 Number of Components and Initialisation

The number of components *k* was set equal to the number of diagnostic classes (*k* = 2 for binary and *k* = 3 for ternary tasks) to encourage a one-to-one correspondence between sources and tissue types; clear class clustering was observed in weight space, with some overlap for spectrally similar classes (Figure 1). cNMF was run for at most 5000 iterations per initialisation, minimising the Frobenius norm ||**V** *−* **WH**|| *_F_*, and repeated over multiple random initialisations to mitigate local optima. One run per (experiment, echo time) was retained, selected once by visual inspection of the sources for correspondence to known tumour biochemistry, because the representation must support physically meaningful spectra rather than merely minimise numerical residual.

**Fig. 1.**
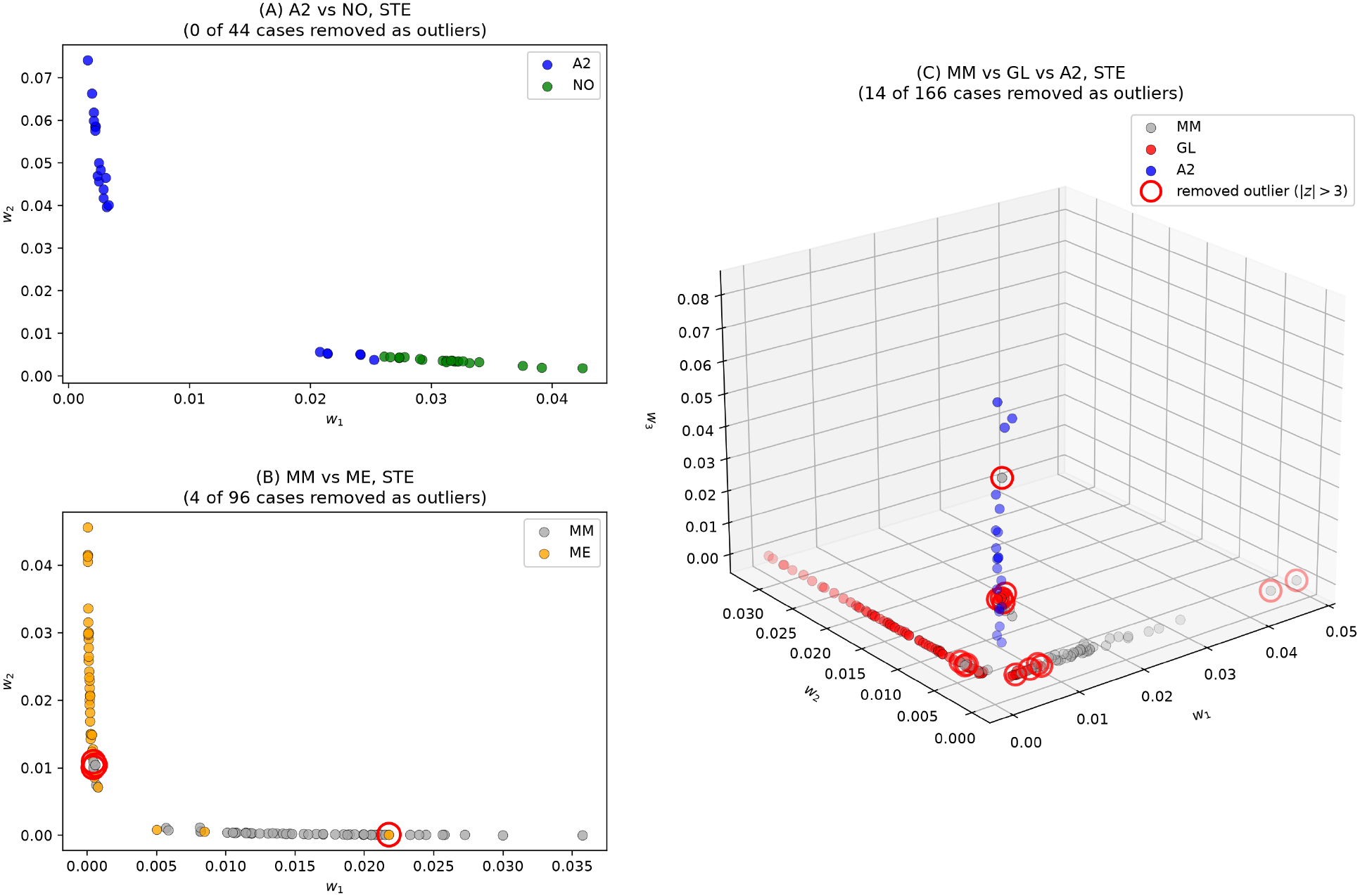
Class separation in the cNMF weight space **H**: (A) a binary task (A2 vs NO, *k* = 2) plotted in its natural 2-dimensional weight space; (B) a ternary task (MM vs ME vs A2, *k* = 3) plotted in its natural 3-dimensional weight space. Each point is one case’s weight vector, coloured by diagnostic class. Red circles mark outliers.

#### 2.2.3 Joint Decomposition

As an alternative to these task-specific decompositions, cNMF was also applied jointly to all five classes for each echo time (*k* = 5), and the weight subset for each of the same six experiments was extracted from the joint weight matrix, allowing a direct comparison under an otherwise identical pipeline (Section 2.4).

#### 2.2.4 Role of the Weight Matrix

The weight matrix **H** produced by cNMF serves as the central representation from which synthetic data are generated. Each column of **H** is a *k*-dimensional weight vector describing how strongly each source component contributes to a given spectrum. Because the source matrix **W** is held fixed after decomposition, any new weight vector **ĥ** sampled from a distribution fitted to **H** can be recombined with **W** to produce a novel synthetic spectrum. The quality and diversity of the resulting synthetic spectra therefore depend directly on how faithfully the generative models capture the true distribution of weights within each class.

The full pipeline from spectral decomposition through to synthetic spectrum reconstruction is illustrated in Figure 2.

**Fig. 2.**
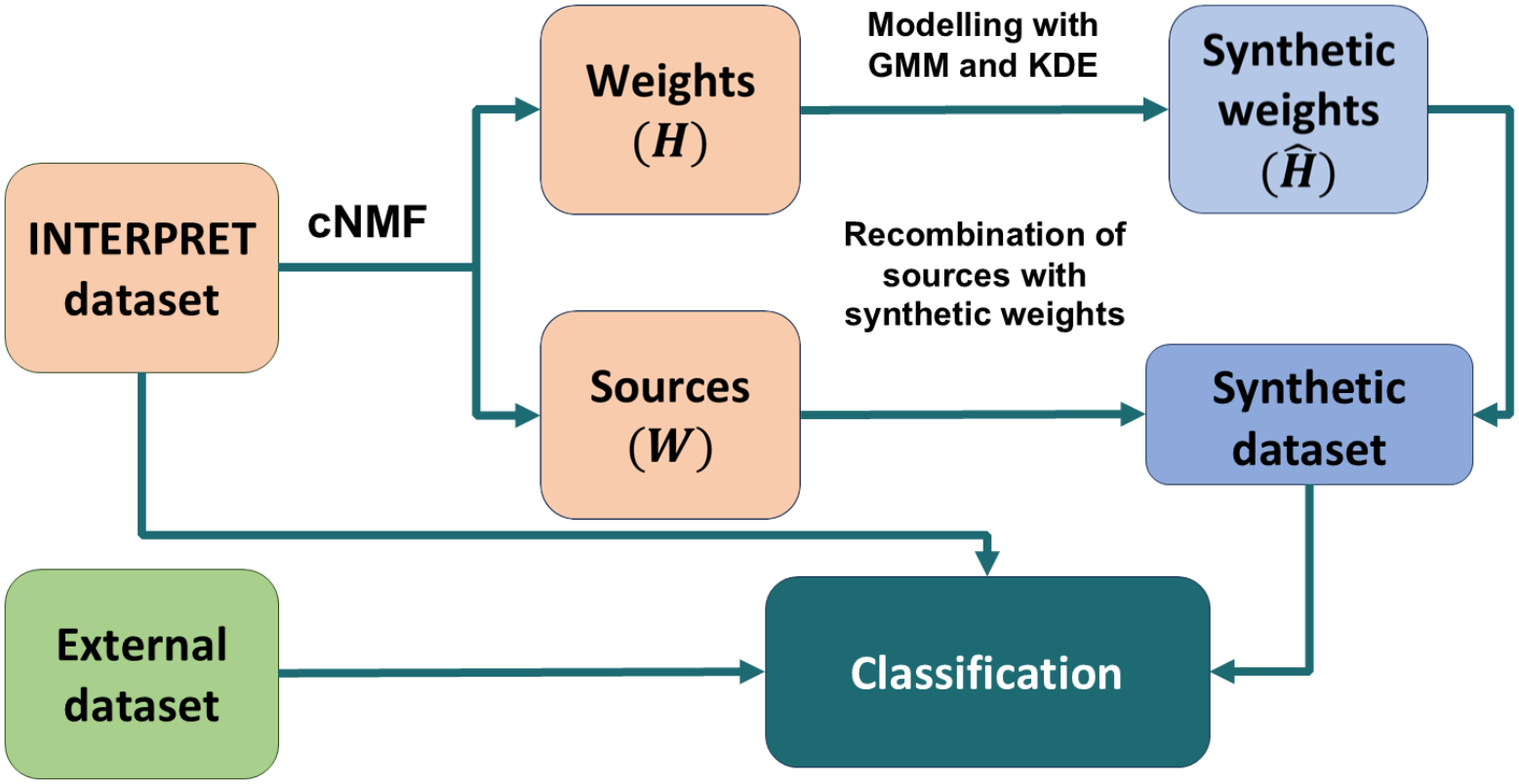
Schematic overview of the proposed pipeline for synthetic MRS spectrum generation. Real spectra from the INTERPRET dataset are decomposed via cNMF into a fixed source matrix **W** and a weight matrix **H**. The weight distributions are segmented by class label, screened for outliers, and modelled using GMM or KDE. Synthetic weight vectors **Ĥ** are sampled from the fitted models and recombined with **W** to reconstruct synthetic spectra **V̂** classification experiments. = **W** *·* **Ĥ**, which are subsequently used in downstream classification experiments.

### 2.3 Generating Plausible Spectra from the Representation

Given the weight representation from Stage 1, generating a synthetic spectrum means drawing a plausible new weight vector for a class and reconstructing it through **W**. This stage therefore has three parts: cleaning the weight distributions of artefacts that would corrupt a fitted model (Section 2.3.1), fitting generative models to those distributions (Section 2.3.2), and sampling and reconstructing spectra from the fitted models (Section 2.3.3). A final part selects, for each task, the generative configuration that reconstructs the real data most faithfully (Section 2.3.4).

#### 2.3.1 Heuristic Outlier Filtering

Prior to distribution modelling, each per-class weight distribution was screened for outliers. Samples whose weight values deviated more than three standard deviations from the class-component mean (*|z| >* 3) were removed. The class distributions in weight space overlap, so a few extreme cases lie well inside the region occupied by another class and would pull a fitted generative model towards it; the weight-space scatter plots show such cases for A2 (Figure 1), and outlying real cases are also visible in the decision-space projection (Figures 7). Removing them is a simple, reproducible way of preventing these observations from biasing the fitted models, at the cost of discarding a small number of atypical but genuine spectra. The number of cases removed per experiment is summarised in Table 3.

**Fig. 3.**
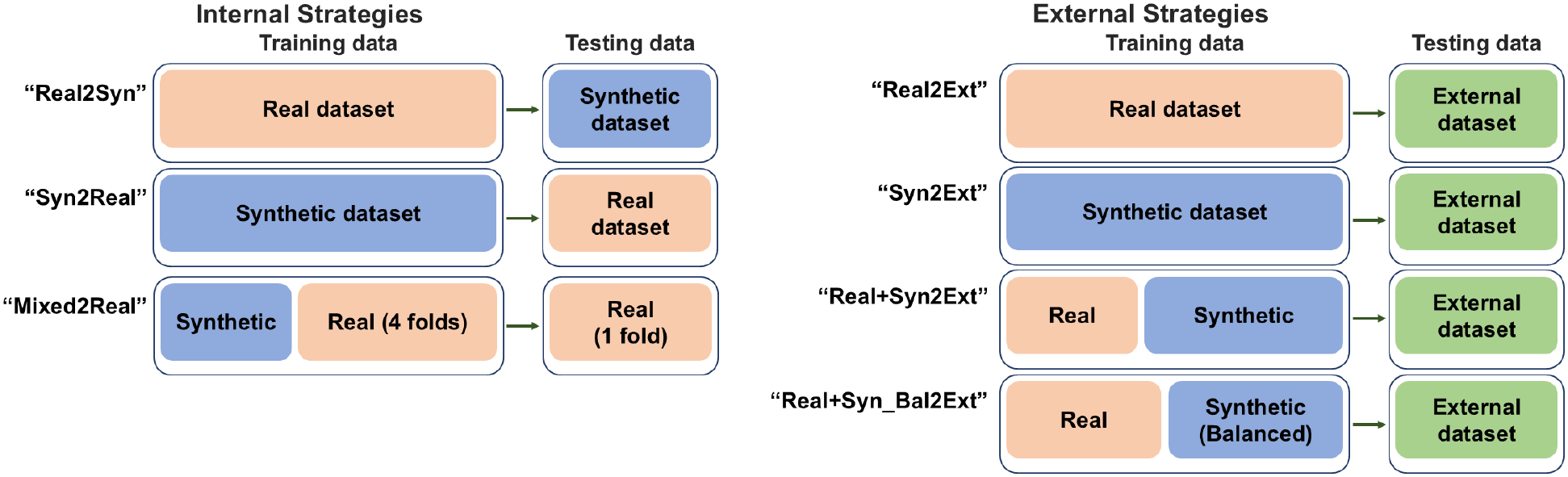
Training and testing data for each classification strategy. Top: internal validation (Real2Syn, Syn2Real, Mixed2Real); bottom: external validation on the independent eTumour cohort (Real2Ext, Syn2Ext, Real+Syn2Ext, Real+Syn Bal.2Ext). Peach: real INTERPRET spectra; blue: synthetic spectra; green: external spectra. Mixed2Real trains on synthetic data plus four real folds and tests on the held-out fold. In Real+Syn Bal.2Ext, synthetic spectra balance the class sizes.

**Fig. 4.**
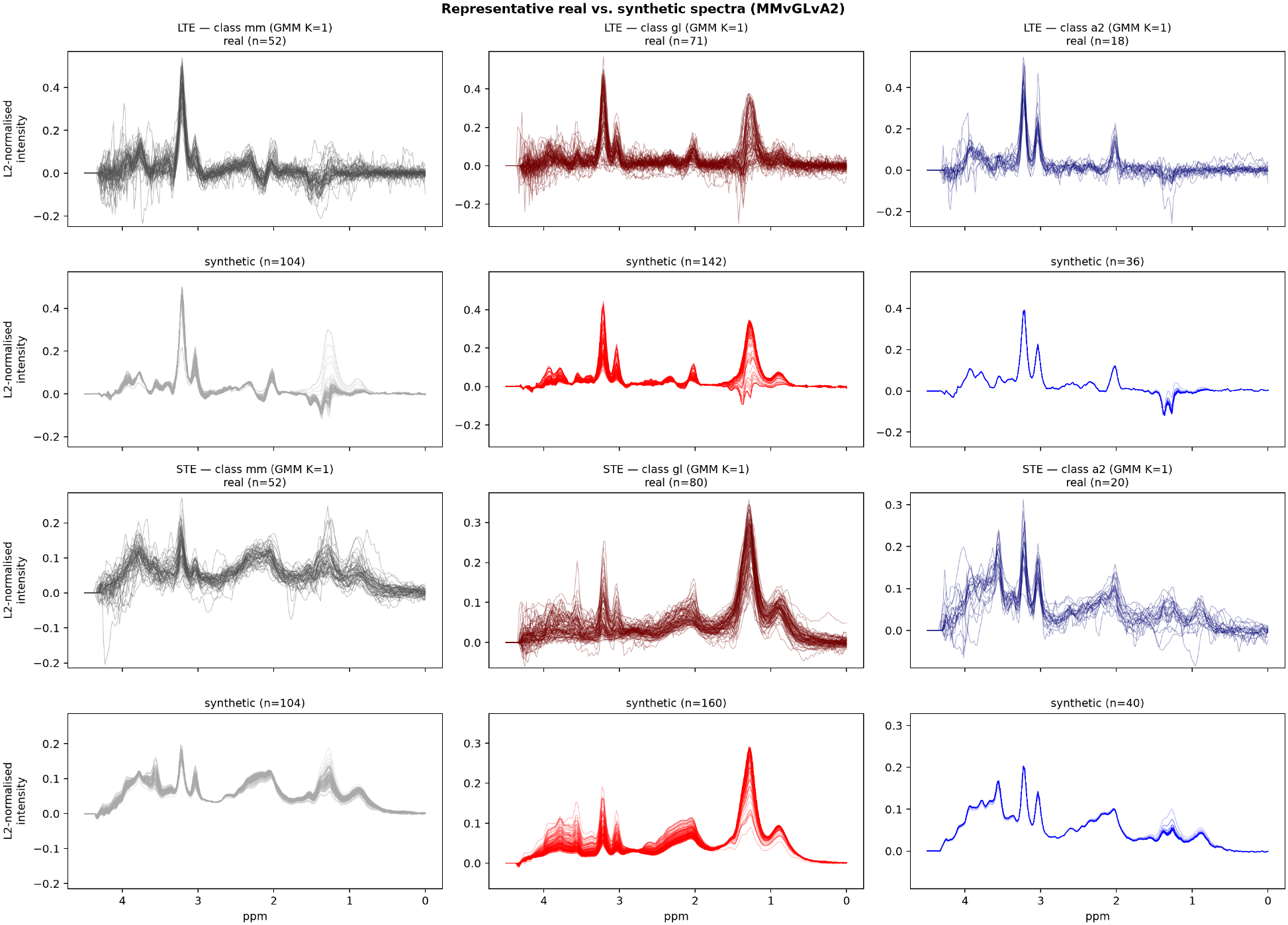
Representative real vs. synthetic spectra for A2 vs NO, at both echo times, using the selected GMM hyperparameter from Table 4. Shaded bands show *±*1 standard deviation. The real band is visibly wider than the synthetic band throughout, consistent with the lower dispersion of synthetic spectra discussed in Section 4.2.

**Fig. 5.**
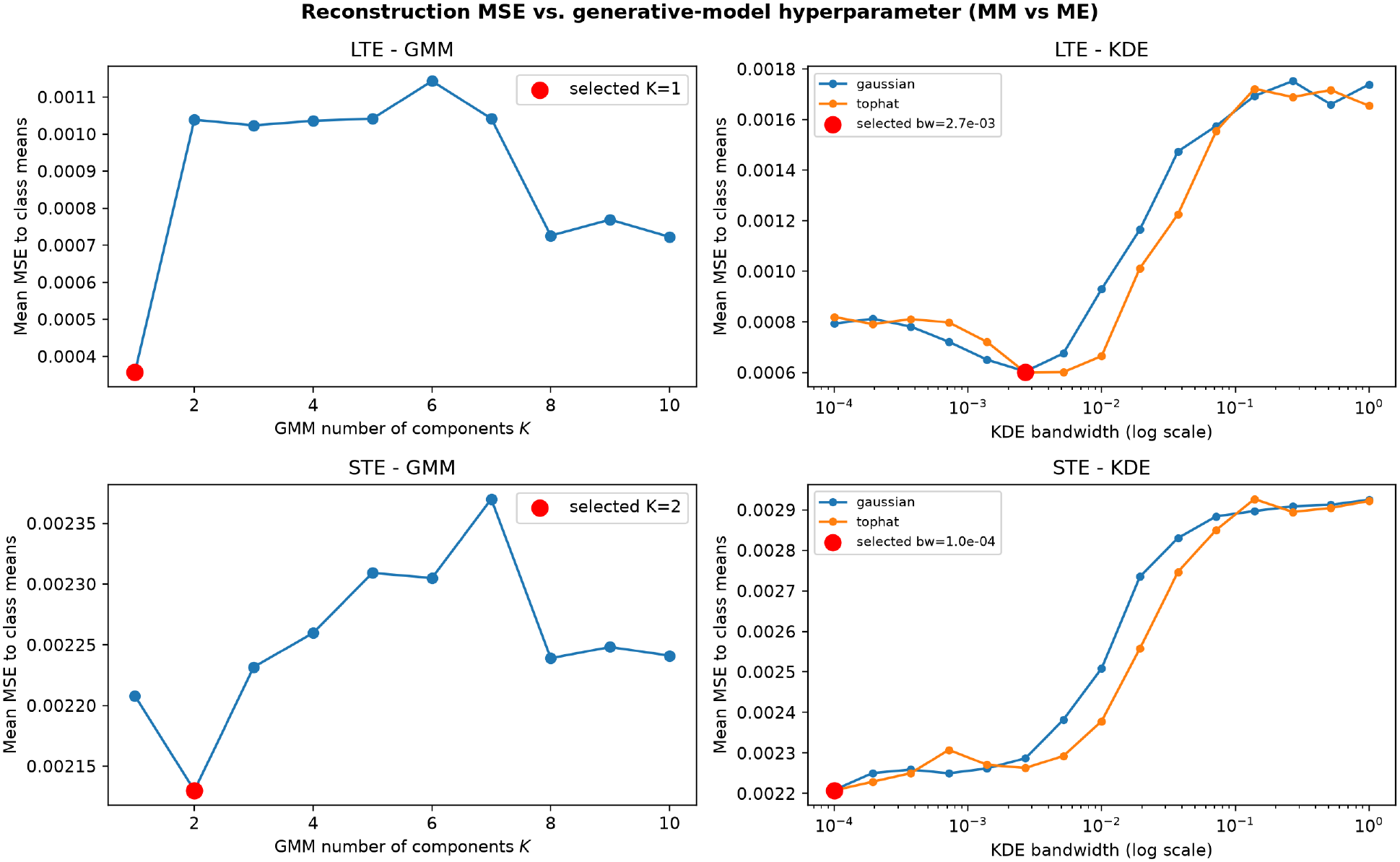
Reconstruction MSE (synthetic spectra vs. real class mean) as a function of the generative-model hyperparameter, for A2 vs NO at both echo times. Left: GMM number of components *K*. Right: KDE bandwidth (log scale), gaussian and tophat kernels. Red markers show the selected configuration reported in Table 4.

**Fig. 6.**
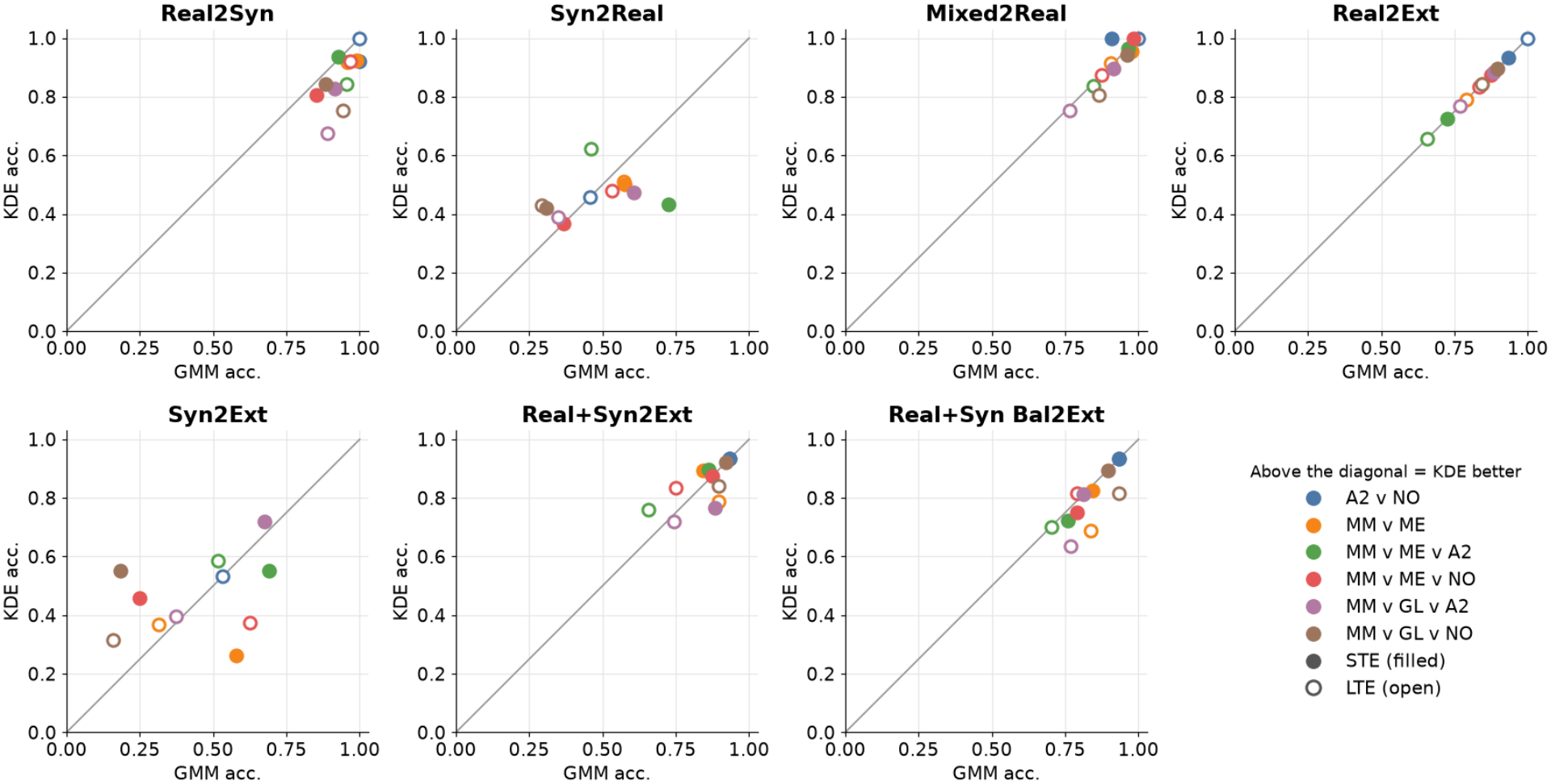
GMM versus KDE accuracy for every task and echo time, one panel per strategy (split decomposition). Points above the diagonal favour KDE. Colour indicates the task; filled markers are STE and open markers are LTE. Real2Ext does not use synthetic data, so it lies on the diagonal.

**Fig. 7.**
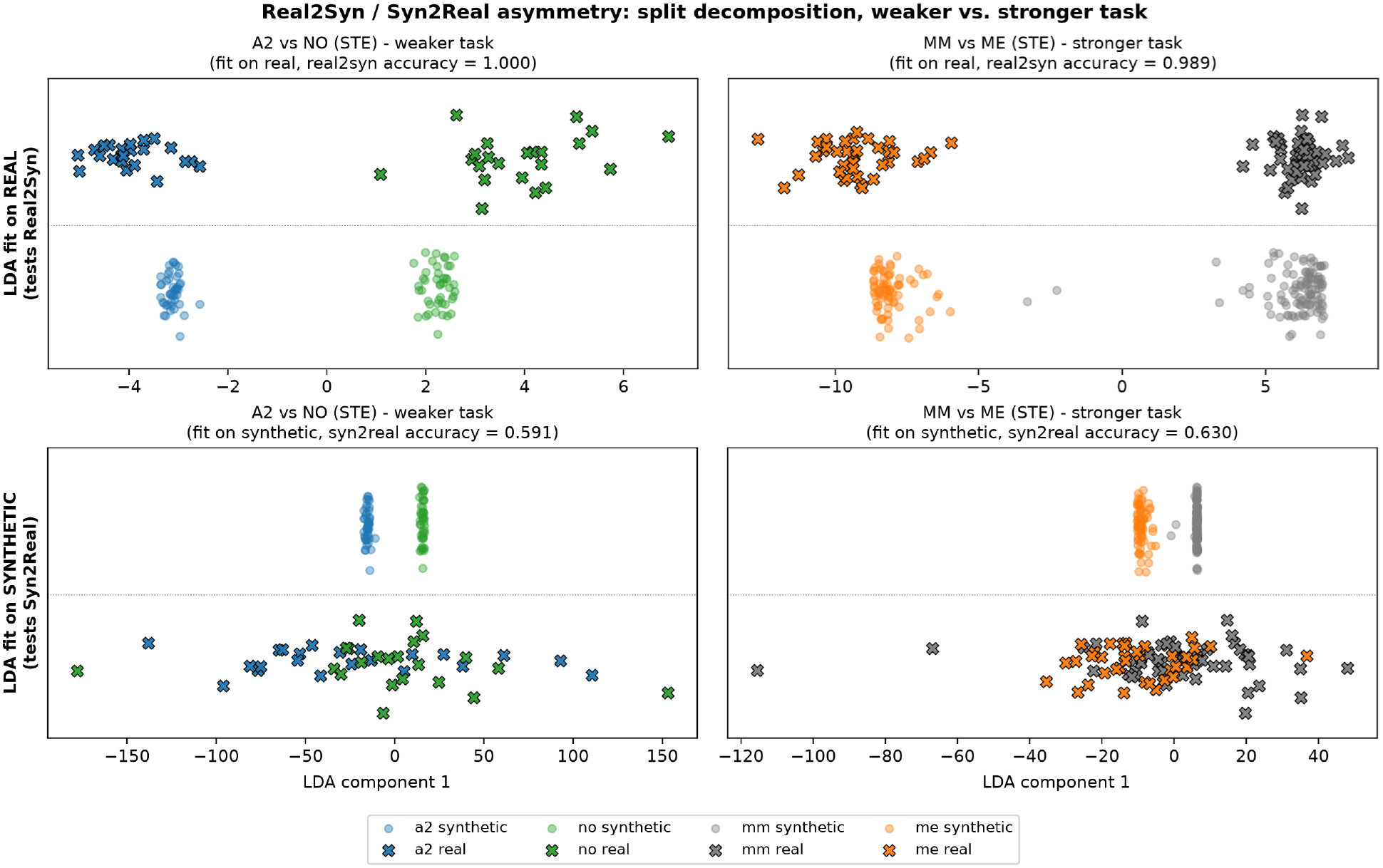
Real and synthetic spectra projected into the fitted LDA decision space, for a weaker task (A2 vs NO) and a stronger one (MM vs ME), task-specific decomposition, STE. Top row: LDA fit on synthetic data (the configuration Syn2Real evaluates). Bottom row: LDA fit on real data (the configuration Real2Syn evaluates). In both tasks, the synthetic points form a tight, well-separated cluster while the real points overlap substantially when projected through the synthetic-fitted boundary (top), but not when the boundary is fitted on real data instead (bottom) — illustrating that the asymmetry is a property of the synthetic distribution’s spread, not of task difficulty.

**Table 3.** Number of cases removed as weight-space outliers (*|z| >* 3 within class) per experiment and echo time, out of the number of cases available before removal (in brackets).

| **Experiment** | **# cases LTE** | **# cases STE** |
|---|---|---|
| A2 vs NO | 0 (35) | 0 (44) |
| MM vs ME | 2 (86) | 4 (96) |
| MM vs ME vs A2 | 8 (106) | 9 (118) |
| MM vs ME vs NO | 5 (101) | 9 (118) |
| MM vs GL vs A2 | 12 (153) | 14 (166) |
| MM vs GL vs NO | 8 (148) | 11 (166) |
| Joint (5 classes) | 10 (199) | 14 (226) |

#### 2.3.2 Generative Modelling of Weight Distributions

The filtered weight matrix **H**, segmented by class label, was used to fit generative models independently for each class, for both the split and joint decompositions. Two complementary modelling strategies were employed and retained in parallel through the remainder of the pipeline (i.e. synthetic data generated by each was carried forward to classification separately, rather than one being selected over the other).

##### Gaussian Mixture Models

GMMs represent the underlying distribution as a weighted sum of *K* Gaussian components, each parametrised by its own mean vector, covariance matrix, and mixture weight. Models were fitted using the Expectation-Maximisation (EM) algorithm with full covariance matrices, as implemented in scikit-learn [27]. The number of mixture components *K* was treated as a hyperparameter and varied from 1 (reducing to a single Gaussian fit) to 10, enabling assessment of how model complexity affects synthetic data quality.

##### Kernel Density Estimation

KDE is a non-parametric approach that estimates the underlying probability density function by placing a kernel over each observed data point. Two kernel types were evaluated: Gaussian and tophat. The bandwidth parameter *h*, which governs the smoothness of the estimated density, was varied on a logarithmic scale from 10*^−^*^4^ to 1. KDE was implemented using the KernelDensity estimator from scikit-learn [27].

#### 2.3.3 Synthetic Weight Sampling and Spectrum Reconstruction

For each class, synthetic weight vectors were sampled from the fitted generative model, generating twice the number of original (post-filtering) samples for that class (synthetic-to-real ratio of 2:1). Because the cNMF weight space is non-negative, sampling was performed by rejection: samples with any negative component were discarded, up to a maximum of 20 sampling attempts. Rejection depends only on the sign of the sampled weights: no signal-to-noise-based criterion is applied to synthetic spectra. The proportion of rejected samples and the number of times the attempt limit was reached were recorded for every class and hyperparameter setting (Table 5). Where this attempt limit was reached before the target sample count was met, remaining synthetic weights were drawn from the fitted model and clipped to a small positive floor of *ε* = 10*^−^*^10^ in place of rejection:

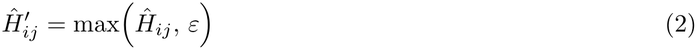

Synthetic spectra were then reconstructed by combining the sampled weights with the fixed source matrix and L2-normalising each row:

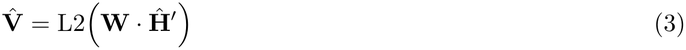

#### 2.3.4 Selecting the Generative Configuration by Reconstruction Fidelity

For each experiment, echo time and model family, the hyperparameter (GMM components, or KDE bandwidth and kernel) was chosen by minimising the mean squared error (MSE) between the synthetic spectra and a reference class mean, with *n* frequency bins, synthetic intensity *x_i_* and real class mean *y_i_*:

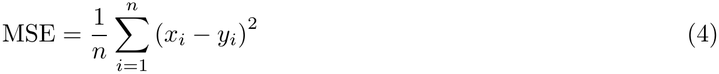

The class mean was computed once per class from all L2-normalised real spectra pooled across split experiments, so that one reference was used throughout. The MSE was averaged across classes, synthetic spectra were then reconstructed only at the optimum, and GMM and KDE settings were retained independently, with no selection between the two families. As a reference level, the same MSE was computed for the cNMF reconstruction of the cleaned real weights; this baseline is not a lower bound, since a generator concentrated near the class centre can score lower than the real weights. For the joint decomposition the sweep was run once for all five classes and the hyperparameter was re-selected by the same criterion over only the classes of each experiment.

For the joint decomposition, synthetic weights were generated once for all five classes and the hyperparameter grid was swept once. When the resulting synthetic data were used for a given two- or three-class experiment, the hyperparameter was re-selected by the same MSE criterion averaged over only the classes of that experiment, so that a model complexity chosen to fit unrelated classes was not imposed on a task with a smaller per-class sample. In one configuration (joint decomposition, GMM, A2 vs NO at STE) the number of GMM components was instead fixed manually to *K* = 1 rather than selected automatically. This is a deviation from the otherwise automatic selection procedure and is noted as a limitation in the Discussion.

### 2.4 Classification Pipeline

The final stage tests the study’s central hypothesis. The key methodological commitment here is that internal performance alone cannot answer it: a generative model that merely memorises the training distribution can inflate cross-validated accuracy while offering no benefit, or even harm, on data from an unseen centre. Our evaluation is therefore built in two tiers, an internal tier that validates the generative model’s behaviour, and an external tier, against a fully independent cohort, that constitutes the actual test of clinical value. Figure 2 summarises the pipeline.

#### 2.4.1 Internal Validation Strategies

Three complementary internal evaluation strategies were applied to each experimental configuration:

- **Real2Syn** — the classifier is trained on real spectra and evaluated on synthetic spectra, assessing whether synthetic data occupies a compatible decision space.
- **Syn2Real** — the classifier is trained exclusively on synthetic spectra and evaluated on real spectra, testing whether synthetic data can substitute for real training data.
- **Mixed2Real** — the classifier is trained on real spectra combined with all synthetic spectra for that experiment, and evaluated on withheld real spectra, simulating a data augmentation scenario.

Real2Syn and Syn2Real involve training and test sets drawn from inherently separate populations (real vs. synthetic) and were therefore each evaluated as a single run, with the standard deviation estimated by bootstrap resampling (1000 resamples) of the test predictions. Mixed2Real was evaluated using 5-fold stratified cross-validation on the real data: within each fold, the held-out real partition served as the test set, while the training set comprised the remaining real folds plus the full synthetic set for that experiment; results were averaged across the five folds. The synthetic spectra used in Mixed2Real, and in Real2Syn and Syn2Real, were generated from all real spectra of the experiment, including those that constitute the held-out real partition in a given fold; the internal tier therefore does not provide a leakage-free estimate of the benefit of augmentation, which is the reason the external tier is treated as the primary test (see Discussion).

#### 2.4.2 External Validation Strategies

The external tier tests generalisation against the independent cohort of Section 2.1.2, and is the primary measure of clinical applicability. Four strategies isolate the contribution of synthetic data against the real-data-only baseline:

- **Real2Ext** — trained on internal real spectra only (baseline).
- **Syn2Ext** — trained on synthetic spectra only.
- **Real Syn2Ext** — trained on internal real spectra plus the full synthetic set.
- **Real Syn Bal2Ext** — trained on real spectra augmented with synthetic spectra sampled to bring each minority class up to the size of the largest real class (class-balancing).

Real2Ext, Syn2Ext and Real Syn2Ext, involve no random sampling and were each evaluated as a single deterministic run, with the external test set bootstrapped to obtain the standard deviation. Real Syn Bal2Ext depends on a random draw of synthetic spectra for class-balancing and was therefore averaged over 10 random seeds; feature selection for this strategy was fixed using a single reference seed to keep the selected feature set consistent across the 10 evaluation runs.

#### 2.4.3 Feature Selection

Feature selection was performed using the CfsSubsetEval criterion in Weka [30], coupled with GreedyStep-wise search operating in backwards elimination mode, approximating Sequential Floating Forward Selection (SFFS). This approach identifies a minimal, non-redundant subset of spectral indices with high predictive utility. A maximum of ten features were retained. This limit was chosen as a reasonable number based on the ranges used in previous MRS classification studies. Features were selected on the training data of the strategy being evaluated (real, synthetic, or their union): within each training partition for Mixed2Real, on the real training set for Real2Syn, and once on the full training set for Real2Ext, Real Syn2Ext, Syn2Ext and Real Syn Bal2Ext (the latter using the reference seed). The exception is Syn2Real, whose features were selected on the real spectra, which are also its test set. This real-basis choice lets the test data influence the feature set and makes the Syn2Real estimate optimistic; we therefore also report the estimate obtained when features are selected on the synthetic training set. When CFS retained fewer than two features, the first two spectral bins were used so that a classifier could be fitted. This occurred twice, both in the split KDE decomposition of A2 vs NO at STE (Syn2Ext, and one Mixed2Real fold); in both cases these bins were constant in the synthetic spectra and the classifier could not be fitted, so the two corresponding results are missing.

#### 2.4.4 Classifier

Classification was performed using Linear Discriminant Analysis (LDA), which identifies a linear projection of the feature space that maximises the ratio of between-class to within-class scatter. LDA was selected for its established performance in MRS-based tumour classification [25], its robustness in small-sample settings, and its interpretability, properties well suited to the goal of evaluating synthetic data contributions rather than maximising predictive performance through model complexity.

##### Performance Metrics

Performance was quantified by accuracy and the Balanced Error Rate (BER), 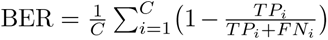 (equivalently one minus balanced accuracy), which is invariant to class imbalance. Accuracy is the value of a single fitted model for Real2Syn, Syn2Real, Real2Ext, Real+Syn2Ext and Syn2Ext, the mean over five folds for Mixed2Real, and the mean over ten balancing seeds for Real+Syn Bal.2Ext. The accompanying standard deviation therefore has a different meaning for each: cross-validation variability for Mixed2Real, bootstrap variability over the test set (*B* = 1000, model fitted once) for the single-fit strategies, and seed variability for Real+Syn Bal.2Ext. These are not directly comparable.

##### Statistical Comparison of Augmentation Strategies

To assess whether synthetic data augmentation improved external generalisation relative to the Real2Ext baseline, paired differences in accuracy were computed across the twelve (experiment *×* echo time) settings for each augmentation strategy, decomposition mode (split/joint), and generative model family (GMM/KDE). Statistical significance of the mean paired difference was assessed using a two-sided Wilcoxon signed-rank test, with zero differences discarded. Effect size was quantified using the rank-biserial correlation *r*, and 95% confidence intervals on the mean difference were obtained by bootstrap resampling (10,000 resamples). The same procedure was used to compare the joint with the split decomposition. No correction for multiple comparisons was applied to the reported *p*-values; where relevant we state which results survive a Bonferroni correction.

## 3 Results

### 3.1 Reconstruction Fidelity and Hyperparameter Selection

Table 4 reports the generative configuration selected for each task-specific decomposition (Section 2.3.4), together with the mean MSE between the synthetic spectra and the real class means. As a reference level, the table also gives the MSE, to the same class means, of the cNMF reconstruction of the *real* weights before any resampling.

**Table 4.** Optimal generative-model hyperparameter and the corresponding mean MSE between synthetic spectra and the real class mean (averaged across classes), per experiment, echo time and model family, for the task-specific decompositions; the joint five-class decomposition is shown separately. The last column is the baseline: the MSE to the same class means (same L2 convention) of the cNMF reconstruction of the *real* weights before any resampling. KDE parameters are (bandwidth, kernel).

| Experiment | Echo | GMM *K* | GMM MSE | KDE (bw, kernel) | KDE MSE | Baseline (real weights) |
|---|---|---|---|---|---|---|
| A2 vs NO | LTE | 5 | 0.000173 | (1.0 × 10<sup>−4</sup>, tophat) | 0.000174 | 0.000174 |
| A2 vs NO | STE | 2 | 0.001450 | (7.2 × 10<sup>−4</sup>, Gaussian) | 0.001464 | 0.001480 |
| MM vs ME | LTE | 1 | 0.000357 | (2.7 × 10<sup>−3</sup>, tophat) | 0.000600 | 0.000782 |
| MM vs ME | STE | 2 | 0.002130 | (1.0 × 10<sup>−4</sup>, tophat) | 0.002208 | 0.002188 |
| MM vs ME vs A2 | LTE | 1 | 0.000286 | (5.2 × 10<sup>−3</sup>, tophat) | 0.000479 | 0.000554 |
| MM vs ME vs A2 | STE | 1 | 0.001996 | (3.7 × 10<sup>−4</sup>, tophat) | 0.002046 | 0.002047 |
| MM vs ME vs NO | LTE | 1 | 0.000330 | (7.2 × 10<sup>−4</sup>, Gaussian) | 0.000517 | 0.000592 |
| MM vs ME vs NO | STE | 7 | 0.001939 | (1.0 × 10<sup>−4</sup>, tophat) | 0.001992 | 0.001975 |
| MM vs GL vs A2 | LTE | 1 | 0.000525 | (5.2 × 10<sup>−3</sup>, tophat) | 0.000618 | 0.000833 |
| MM vs GL vs A2 | STE | 1 | 0.002224 | (5.2 × 10<sup>−3</sup>, tophat) | 0.002291 | 0.002341 |
| MM vs GL vs NO | LTE | 1 | 0.000453 | (5.2 × 10<sup>−3</sup>, tophat) | 0.000651 | 0.000746 |
| MM vs GL vs NO | STE | 3 | 0.002038 | (5.2 × 10<sup>−3</sup>, tophat) | 0.002082 | 0.002124 |
| *Joint decomposition (all five classes, k* = 5*)* |  |  |  |  |  |  |
| Joint | LTE | 2 | 0.000489 | (3.7 × 10<sup>−2</sup>, gaussian) | 0.000572 | 0.000681 |
| Joint | STE | 4 | 0.001903 | (1.0 × 10<sup>−2</sup>, tophat) | 0.002043 | 0.001987 |

Reconstruction MSE was four to eight times higher at STE (1.4–2.3 *×* 10*^−^*^3^) than at LTE (1.7 *×* 10*^−^*^4^– 6.5 *×* 10*^−^*^4^) for every experiment, so MSE values are only comparable within an echo time. Within an echo time, the GMM optimum was at least as good as the KDE optimum in all 14 configurations (12 task-specific, 2 joint), so there was no configuration in which KDE gave the lower MSE. The margin was small at STE (KDE at most 4% higher for task-specific decompositions) and larger at LTE, where the KDE optimum was up to 1.7 times higher (e.g. MM vs ME, LTE: 3.6 *×* 10*^−^*^4^ for GMM against 6.0 *×* 10*^−^*^4^ for KDE).

The selected model complexity was low. A single-component GMM (*K* = 1, i.e. a single full-covariance Gaussian per class) was selected in 7 of the 12 task-specific configurations, including five of the six LTE experiments; the remaining optima were *K* = 2–3 (three configurations: A2 vs NO STE, MM vs ME STE, MM vs GL vs NO STE), *K* = 5 (A2 vs NO, LTE) and *K* = 7 (MM vs ME vs NO, STE). The selected KDE bandwidths were correspondingly small (10*^−^*^4^ to 5.2 *×* 10*^−^*^3^ on a grid spanning 10*^−^*^4^ to 1) and the tophat kernel was selected in 10 of 12 configurations. For the joint decomposition the optimum was *K* = 2 (LTE) and *K* = 4 (STE) for the GMM, with KDE bandwidths of 3.7 *×* 10*^−^*^2^ and 10*^−^*^2^ (Table 4).

Notably, at the selected GMM setting the synthetic spectra were at least as close to the real class means as the reconstruction of the real weights themselves, in all 12 task-specific and both joint configurations (ratio of synthetic to baseline MSE between 0.46 and 1.00); for KDE this held in 10 of 14 configurations (ratio 0.74–1.03). The real-weight reconstruction is therefore not a lower bound on the MSE that synthetic spectra can attain. The gap was largest at LTE for *K* = 1 models; for MM vs ME (LTE) the MSE of the synthetic spectra was 3.6 *×* 10*^−^*^4^ against 7.8 *×* 10*^−^*^4^ for the real-weight reconstruction, and for MM vs ME vs A2 (LTE) 2.9 *×* 10*^−^*^4^ against 5.5 *×* 10*^−^*^4^. Synthetic spectra that lie closer to the class mean than the real spectra do are, on average, less dispersed about that mean than the real data; we return to this in the Discussion.

#### Rejection sampling

Synthetic weight vectors were rejected when any component was non-positive (Section 2.3.3); no signal- to-noise-based rejection is applied at this stage. Table 5 gives the per-class rejection rates at the selected hyperparameters. On average 54% (GMM) and 53% (KDE) of the sampled weight vectors were rejected, and the rate was highest for the single-component GMMs (62% on average, range 41–93%). Rejection was not uniform across classes: for the task-specific decompositions it was highest for GL (66% GMM, 73% KDE) and MM (58%, 50%) and lowest for A2 (39%, 45%), and for the joint decomposition it was highest for NO (72% GMM, 78% KDE, against 52–68% for the other classes). The 20-attempt cap, after which remaining weights were clipped to *ε*, was reached once at the selected hyperparameters (MM vs ME vs NO, LTE, split GMM *K* = 1, class NO: 93% rejected, 4 of 30 weight vectors clipped), and in 32 of the 1680 class–hyperparameter combinations of the full sweep (25 GMM, 7 KDE).

### 3.2 Internal Validation

Table 6 summarises performance by strategy, and Table 7 gives the internal results per experiment and echo time, pooled over the four decomposition–model configurations (results for every configuration are provided in the supplementary tables). The three internal strategies separated clearly.

**Table 5.**
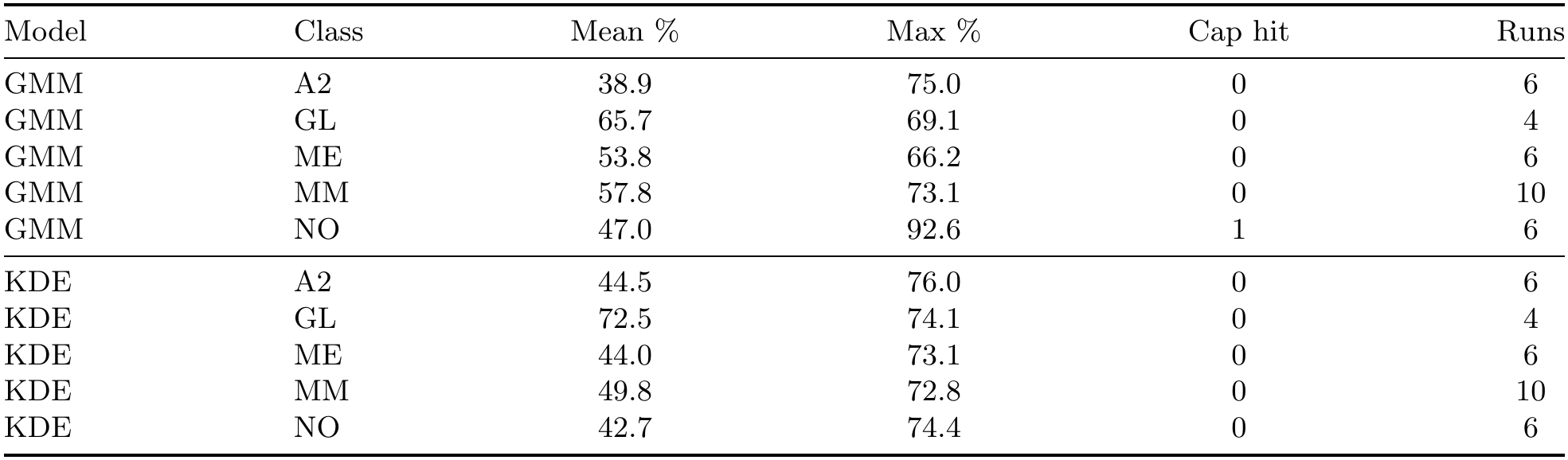
Rejection sampling at the selected hyperparameter (task-specific decompositions): mean and maximum percentage of sampled weight vectors rejected for having a non-positive component, per class, over the runs in which the class appears, and the number of runs in which the 20-attempt cap was reached (clipping fallback used).

**Table 6.** Mean accuracy and balanced error rate (BER) across the twelve (experiment *×* echo time) settings, by evaluation strategy, decomposition and generative model.

|  |  | Split |  | Joint |  |
|---|---|---|---|---|---|
| Strategy | Metric | GMM | KDE | GMM | KDE |
| Real2Syn | Acc. | 0.940 | 0.864 | 0.876 | 0.815 |
|  | BER | 0.048 | 0.119 | 0.128 | 0.196 |
| Syn2Real | Acc. | 0.534 | 0.472 | 0.590 | 0.615 |
|  | BER | 0.439 | 0.500 | 0.379 | 0.378 |
| Mixed2Real | Acc. | 0.914 | 0.911 | 0.884 | 0.902 |
|  | BER | 0.077 | 0.091 | 0.108 | 0.092 |
| Real2Ext | Acc. | 0.837 | 0.837 | 0.839 | 0.839 |
|  | BER | 0.159 | 0.159 | 0.157 | 0.157 |
| Real+Syn2Ext | Acc. | 0.849 | 0.847 | 0.815 | 0.840 |
|  | BER | 0.136 | 0.153 | 0.184 | 0.158 |
| Syn2Ext | Acc. | 0.442 | 0.466 | 0.432 | 0.557 |
|  | BER | 0.531 | 0.522 | 0.555 | 0.422 |
| Real+Syn Bal.2Ext | Acc. | 0.833 | 0.795 | 0.822 | 0.820 |
|  | BER | 0.151 | 0.186 | 0.169 | 0.166 |

**Table 7.** Internal classification accuracy by experiment and echo time, pooled over the four decomposition (split, joint) *×* generative model (GMM, KDE) configurations: mean of the four single-run accuracies *±* root-mean-square of their standard deviations. The *±* is a standard deviation whose meaning depends on the strategy: Mixed2Real, 5-fold cross-validation variability; Real2Syn, Syn2Real, Real2Ext, Real+Syn2Ext and Syn2Ext, bootstrap variability over the test set (*B* = 1000, single fitted model); Real+Syn Bal.2Ext, variability over 10 balancing seeds. Asterisks mark a significant difference from the Real2Syn baseline (two-sided Fisher exact test on the correct/incorrect counts of matching configurations): *^∗^ p <* 0.05, *^∗∗^ p <* 0.01, *^∗∗∗^ p <* 0.001. Each cell reports the least significant *p* over its four configurations, so the difference holds in all four; *p*-values are uncorrected for multiple comparisons. Cell shading shows the accuracy on a diverging scale (red below 0.5, white at 0.5, green above 0.5).

| Experiment | Echo | Real2Syn | Syn2Real | Mixed2Real |
|---|---|---|---|---|
| A2 vs NO | LTE | 1.000 ± 0.000 | 0.380 ± 0.082<sup>***</sup> | 0.967 ± 0.078 |
|  | STE | 0.819 ± 0.040 | 0.612 ± 0.074 | 0.924 ± 0.076 |
| MM vs ME | LTE | 0.905 ± 0.023 | 0.640 ± 0.052<sup>*</sup> | 0.911 ± 0.055 |
|  | STE | 0.944 ± 0.017 | 0.604 ± 0.050<sup>***</sup> | 0.960 ± 0.045 |
| MM vs ME vs A2 | LTE | 0.850 ± 0.025 | 0.518 ± 0.050<sup>**</sup> | 0.847 ± 0.063 |
|  | STE | 0.907 ± 0.019 | 0.634 ± 0.044 | 0.935 ± 0.048 |
| MM vs ME vs NO | LTE | 0.904 ± 0.021 | 0.463 ± 0.049<sup>***</sup> | 0.878 ± 0.059 |
|  | STE | 0.853 ± 0.024 | 0.409 ± 0.047<sup>***</sup> | 0.971 ± 0.034 |
| MM vs GL vs A2 | LTE | 0.720 ± 0.025 | 0.368 ± 0.041<sup>***</sup> | 0.766 ± 0.042 |
|  | STE | 0.859 ± 0.020 | 0.512 ± 0.040<sup>***</sup> | 0.910 ± 0.042 |
| MM vs GL vs NO | LTE | 0.831 ± 0.022 | 0.480 ± 0.040<sup>***</sup> | 0.829 ± 0.102 |
|  | STE | 0.894 ± 0.018 | 0.411 ± 0.038<sup>***</sup> | 0.939 ± 0.043 |

Mixed2Real, in which the classifier is trained on real spectra plus the full synthetic set and evaluated by 5-fold cross-validation on held-out real spectra, achieved a mean accuracy of 0.903 (range 0.752–1.000; mean BER 0.092). Real2Syn, in which a classifier trained on real spectra is evaluated on synthetic spectra, achieved a mean accuracy of 0.874 (range 0.610–1.000; mean BER 0.123), indicating that synthetic spectra are for the most part classified correctly by decision boundaries learned from real data. By contrast, Syn2Real, training exclusively on synthetic spectra and evaluating on real spectra, achieved a mean accuracy of only 0.553 (range 0.165–0.901; mean BER 0.424), and was lower than Real2Syn in 45 of the 48 settings. Because the features used for Syn2Real were selected on the real spectra, which are also its test set (Section 2.4.3), this figure is if anything optimistic; when features were selected on the synthetic training set instead, the mean Syn2Real accuracy was 0.500.

This asymmetry is consistent with a domain gap between the synthetic and real spectral distributions: synthetic spectra fall inside regions of feature space that real-trained classifiers assign to the correct class, but they do not span the real distribution well enough for a classifier trained on them alone to generalise back to real data.

The domain gap varied by task and echo time but not systematically by generative model or decomposition. Syn2Real accuracy was highest for MM vs ME (0.789 averaged over echo time, decomposition and model) and A2 vs NO (0.673) and lowest for MM vs ME vs A2 (0.369, close to the three-class chance level of *≈* 0.33), MM vs ME vs NO (0.446) and MM vs GL vs A2 (0.462). Mixed2Real was higher at STE (0.940) than at LTE (0.866), whereas Real2Syn and Syn2Real differed little between echo times (0.879 vs 0.868 and 0.537 vs 0.568). Differences between GMM and KDE were small relative to the setting-to-setting variation (Table 6); mean Syn2Real was 0.562 for GMM and 0.543 for KDE. Syn2Real was higher for the joint (0.602) than for the task-specific decompositions (0.503), the largest such systematic difference among the internal strategies, but the difference went in the opposite direction for Real2Syn (0.845 joint vs 0.902 split) and Mixed2Real (0.893 vs 0.913) and did not carry over to the external comparison (Section 3.4). One configuration (split KDE, A2 vs NO, STE) failed for Syn2Ext, and contributed four rather than five Mixed2Real folds, because feature selection on the synthetic training data retained a single feature and the fallback features (the first two spectral bins) were constant in the synthetic spectra; the internal comparison otherwise covers all 48 settings.

### 3.3 External Validation

Table 8 gives the external accuracy of the four external strategies (Section 2.4.2) per experiment and echo time, pooled over the four decomposition–model configurations (all configurations are given in the supplementary tables), and Table 9 gives the paired comparison of each augmentation strategy with the Real2Ext baseline.

**Table 8.** External classification accuracy on the independent cohort by experiment and echo time, pooled over the four decomposition *×* generative model configurations (mean *±* root-mean-square of the standard deviations). Real2Ext does not use synthetic data, so its four configurations differ only in the real cases retained by outlier removal. The *±* is a standard deviation whose meaning depends on the strategy: Mixed2Real, 5-fold cross-validation variability; Real2Syn, Syn2Real, Real2Ext, Real+Syn2Ext and Syn2Ext, bootstrap variability over the test set (*B* = 1000, single fitted model); Real+Syn Bal.2Ext, variability over 10 balancing seeds. Asterisks mark a significant difference from the Real2Ext baseline (two-sided Fisher exact test on the correct/incorrect counts of matching configurations; conservative, as the same external cases are used by both strategies): *^∗^ p <* 0.05, *^∗∗^ p <* 0.01, *^∗∗∗^ p <* 0.001. Each cell reports the least significant *p* over its four configurations, so the difference holds in all four; *p*-values are uncorrected for multiple comparisons. Cell shading shows the accuracy on a diverging scale (red below 0.5, white at 0.5, green above 0.5).

| Experiment | Echo | Real2Ext | Real+Syn2Ext | Syn2Ext | Real+Syn Bal.2Ext |
|---|---|---|---|---|---|
| A2 vs NO | LTE | 0.967 ± 0.046 | 0.917 ± 0.071 | 0.450 ± 0.128<sup>**</sup> | 0.967 ± 0.000 |
|  | STE | 0.933 ± 0.065 | 0.933 ± 0.065 | 0.622 ± 0.110 | 0.933 ± 0.000 |
| MM vs ME | LTE | 0.842 ± 0.080 | 0.776 ± 0.093 | 0.461 ± 0.112 | 0.745 ± 0.017 |
|  | STE | 0.842 ± 0.082 | 0.855 ± 0.078 | 0.487 ± 0.108 | 0.872 ± 0.018 |
| MM vs ME vs A2 | LTE | 0.690 ± 0.086 | 0.716 ± 0.083 | 0.526 ± 0.092 | 0.723 ± 0.016 |
|  | STE | 0.759 ± 0.080 | 0.836 ± 0.069 | 0.647 ± 0.086 | 0.758 ± 0.012 |
| MM vs ME vs NO | LTE | 0.792 ± 0.082 | 0.781 ± 0.083 | 0.385 ± 0.091 | 0.748 ± 0.013 |
|  | STE | 0.833 ± 0.076 | 0.885 ± 0.066 | 0.365 ± 0.094<sup>*</sup> | 0.792 ± 0.000 |
| MM vs GL vs A2 | LTE | 0.779 ± 0.061 | 0.738 ± 0.066 | 0.372 ± 0.075<sup>***</sup> | 0.695 ± 0.011 |
|  | STE | 0.884 ± 0.048 | 0.837 ± 0.055 | 0.657 ± 0.073 | 0.826 ± 0.007 |
| MM vs GL vs NO | LTE | 0.829 ± 0.061 | 0.849 ± 0.058 | 0.375 ± 0.076<sup>*</sup> | 0.870 ± 0.008 |
|  | STE | 0.908 ± 0.045 | 0.928 ± 0.041 | 0.382 ± 0.069<sup>*</sup> | 0.882 ± 0.000 |

**Table 9.** Paired comparison of each augmentation strategy against the Real2Ext baseline (external accuracy, one pair per experiment *×* echo time setting, *n* = 12 unless a configuration failed): mean paired difference with 95% bootstrap confidence interval, Wilcoxon signed-rank statistic *W*, two-sided *p*-value, and rank-biserial correlation *r* (negative: strategy worse than baseline). The final column gives the number of settings in which the paired difference was positive, negative or zero (+*/ − /* =; strategy minus baseline). *p*-values are uncorrected; with the 12 tests in this table, *p <* 0.004 survives a Bonferroni correction.

| Strategy | Decomp. | Model | Mean Δ [95% CI] | *W* | *p* | *r* | +/ − / = |
|---|---|---|---|---|---|---|---|
| Real+Syn2Ext | Split | GMM | +0.012 [-0.021, +0.048] | 10 | 0.578 | +0.29 | 4/3/5 |
| Real+Syn2Ext | Split | KDE | +0.010 [-0.029, +0.053] | 12 | 0.812 | +0.14 | 4/3/5 |
| Real+Syn2Ext | Joint | GMM | -0.025 [-0.070, +0.013] | 11 | 0.383 | -0.39 | 3/5/4 |
| Real+Syn2Ext | Joint | KDE | +0.000 [-0.041, +0.039] | 16 | 0.844 | +0.11 | 4/4/4 |
| Real+Syn Bal.2Ext | Split | GMM | -0.003 [-0.032, +0.026] | 16 | 0.844 | -0.11 | 4/4/4 |
| Real+Syn Bal.2Ext | Split | KDE | -0.042 [-0.073, -0.013] | 4 | 0.027 | -0.82 | 1/8/3 |
| Real+Syn Bal.2Ext | Joint | GMM | -0.018 [-0.056, +0.017] | 15 | 0.426 | -0.33 | 4/5/3 |
| Real+Syn Bal.2Ext | Joint | KDE | -0.019 [-0.063, +0.023] | 28 | 0.700 | -0.15 | 4/7/1 |
| Syn2Ext | Split | GMM | -0.395 [-0.515, -0.272] | 0 | 0.000 | -1.00 | 0/12/0 |
| Syn2Ext | Split | KDE | -0.362 [-0.449, -0.267] | 0 | 0.001 | -1.00 | 0/11/0 |
| Syn2Ext | Joint | GMM | -0.408 [-0.518, -0.310] | 0 | 0.000 | -1.00 | 0/12/0 |
| Syn2Ext | Joint | KDE | -0.282 [-0.381, -0.175] | 2 | 0.003 | -0.94 | 1/10/1 |

The Real2Ext baseline, trained on real INTERPRET spectra only, achieved a mean external accuracy of 0.838 (BER 0.158), ranging from 0.655 to 1.000 across settings. It was highest for the binary A2 vs NO task (mean 0.950 over echo times) and lowest for MM vs ME vs A2 (0.724), and was higher at STE (0.860) than at LTE (0.816). The *±* values in Table 8 are bootstrap standard deviations over the external test set for every strategy except Real+Syn Bal.2Ext, for which they are standard deviations over the 10 balancing seeds. The bootstrap standard deviations were large relative to the differences between strategies (mean 0.066 for Real2Ext, 0.069 for Real+Syn2Ext a<u>nd 0.092 fo</u>r Syn2Ext), reflecting the small external cohort; they closely matched the binomial expectation 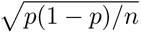 (ratio 0.94–1.06). The seed standard deviations of Real+Syn Bal.2Ext (mean 0.007) reflect only the random draw of synthetic spectra with the external test set held fixed, and are therefore not comparable with the bootstrap values.

Augmenting the real training data with the full synthetic set (Real+Syn2Ext) did not change performance on average: mean accuracy was 0.838, the same as the baseline (difference *<* 0.001). The mean paired difference was +0.012 (split, GMM), +0.010 (split, KDE), *−*0.025 (joint, GMM) and 0.000 (joint, KDE), with each 95% confidence interval spanning zero and *p ≥* 0.38 in every case (Table 9). Individual settings moved in both directions: across the four decomposition–model combinations, augmentation improved accuracy in 3–4 of 12 settings, reduced it in 3–5, and left it unchanged in 4–5 (Table 9). The largest gain was +0.17 (MM vs ME vs A2, STE, split KDE) and the largest loss was *−*0.21 (MM vs ME, LTE, joint GMM), so gains and losses were of comparable magnitude.

Training on synthetic spectra alone (Syn2Ext) was consistently and substantially worse than the baseline. Mean accuracy was 0.474 (BER 0.507), and the paired difference was negative in 45 of the 47 comparisons, with mean differences between *−*0.28 and *−*0.41 across decomposition–model combinations (*p ≤* 0.003 in all four, all significant after Bonferroni correction across the 12 tests of Table 9). Syn2Ext accuracy also varied widely between settings (0.08–0.93), and for the same task it could differ by more than 0.4 between generative models or decompositions (e.g. MM vs ME vs NO at LTE, split versus joint GMM: 0.54 lower under the joint decomposition), which indicates that synthetic-only classifiers are unstable as well as inaccurate. This result mirrors the Syn2Real finding above and indicates that synthetic spectra are not a viable substitute for real training data under cross-cohort evaluation.

The class-balanced strategy (Real+Syn Bal.2Ext) did not help on average: its mean accuracy was 0.817, 0.021 below the baseline. The mean paired difference was *−*0.004 (split, GMM), *−*0.042 (split, KDE), *−*0.018 (joint, GMM) and *−*0.019 (joint, KDE). Only the split KDE difference was nominally significant (*p* = 0.027, 8 of 12 settings worse), and it does not survive correction for the 12 comparisons in Table 9. We had expected the effect of balancing to depend on the degree of class imbalance in the training set. We found no such relationship: across the twelve (experiment *×* echo time) settings, the change in accuracy from balancing was uncorrelated with the ratio of the largest to the smallest training class (Spearman *ρ* = +0.14, *p* = 0.68 for split GMM; *ρ* = *−*0.13, *p* = 0.69 for split KDE). For example, balancing improved MM vs GL vs NO at LTE with split GMM (+0.092, imbalance ratio 4.9) but reduced MM vs GL vs A2 at STE (*−*0.072, ratio 4.0), and had no effect on A2 vs NO at STE (ratio 1.0).

### 3.4 Joint versus Split Decomposition

Table 10 compares the joint (single five-class) decomposition with the task-specific (split) decomposition on external accuracy. No comparison was significant. The mean joint-minus-split difference was +0.003 for the Real2Ext baseline, between *−*0.035 and *−*0.007 for Real+Syn2Ext, between *−*0.010 and +0.057 for Syn2Ext, and between *−*0.012 and +0.026 for Real+Syn Bal.2Ext (all *p ≥* 0.147). The joint decomposition therefore matched the task-specific decomposition within the resolution of this design. Differences within individual settings were nonetheless sometimes large and of either sign. Even the Real2Ext baseline, which depends on the decomposition only through which cases survive outlier removal, differed by up to +0.105 (MM vs ME, LTE) and *−*0.083 (MM vs ME vs NO, both echo times), and Syn2Ext differed between decompositions by as much as +0.42 and *−*0.54 (split GMM versus joint GMM, MM vs GL vs NO at LTE and MM vs ME vs NO at LTE respectively). We therefore interpret the absence of a systematic difference as evidence of comparable average performance rather than of stability.

**Table 10.** Joint minus split external accuracy, paired over the twelve (experiment *×* echo time) settings, by strategy and generative model (Wilcoxon signed-rank, two-sided, uncorrected). The final column gives the number of settings in which the paired difference was positive (joint better), negative (split better) or zero (+*/ − /* =; joint minus split).

| Strategy | Model | Mean Δ | *p* | +/ − / = |
|---|---|---|---|---|
| Real2Ext | GMM | +0.003 | 0.883 | 5/4/3 |
| Real2Ext | KDE | +0.003 | 0.883 | 5/4/3 |
| Real+Syn2Ext | GMM | -0.035 | 0.203 | 2/6/4 |
| Real+Syn2Ext | KDE | -0.007 | 0.643 | 4/6/2 |
| Syn2Ext | GMM | -0.010 | 0.966 | 5/6/1 |
| Syn2Ext | KDE | +0.057 | 0.320 | 7/4/0 |
| Real+Syn Bal.2Ext | GMM | -0.012 | 0.625 | 5/5/2 |
| Real+Syn Bal.2Ext | KDE | +0.026 | 0.147 | 8/3/1 |

### 3.5 Statistical Comparison of Augmentation Strategies

Paired differences in external accuracy between each augmentation strategy and the Real2Ext baseline were assessed with a two-sided Wilcoxon signed-rank test over the twelve (experiment *×* echo time) settings, separately for each decomposition mode and generative model (Table 9), with effect sizes given as the rank-biserial correlation *r* and 95% confidence intervals on the mean difference from 10,000 bootstrap resamples.

Neither augmentation strategy that retained real data (Real+Syn2Ext, Real+Syn Bal.2Ext) produced a significant improvement in any of the eight comparisons. Real+Syn2Ext showed no significant change in any of its four comparisons, whereas Real+Syn Bal.2Ext was nominally worse in one (split decomposition, KDE: *−*0.042, *p* = 0.027), a difference that did not survive correction for multiple comparisons. The 95% confidence intervals on the mean difference had half-widths of about 0.03–0.04, and their upper limits were at most +0.053 in all eight comparisons, so there is no evidence that augmentation caused harm in general, but also no evidence of benefit. The only nominally significant result for these strategies was a *decrease* for Real+Syn Bal.2Ext with split KDE (*p* = 0.027, uncorrected). Syn2Ext was significantly worse than the baseline in all four decomposition–model combinations, with very large negative effect sizes (*r ≤ −*0.94).

The confidence intervals for the class-balanced strategy were narrower than those for Real+Syn2Ext in three of the four decomposition–model combinations (widths 0.058–0.073 against 0.069–0.083), consistent with its more stable accuracy across settings, and the one nominally significant result for these strategies was a decrease.

## 4 Discussion

### 4.1 Principal Findings

This study asked whether synthetic MR spectra, generated by cNMF decomposition and resampling of the resulting weight distributions, can improve the external generalisation of brain tumour classifiers. Across six diagnostic tasks, two echo times, two decomposition strategies and two generative model families, the answer was no. Augmenting real training data with synthetic spectra did not significantly change accuracy on an independent multi-centre cohort (Real+Syn2Ext: mean paired differences between *−*0.03 and +0.01; all *p ≥* 0.38). Class-balanced augmentation did not help either; it was nominally worse in one of four comparisons (*−*0.042, *p* = 0.027, not significant after correction) and indistinguishable from the baseline in the other three. The lack of harm is itself informative: synthetic spectra can be added to real training data at no measurable cost, even though they bring no measurable benefit. Training on synthetic spectra alone was significantly worse than training on real data (mean external accuracy 0.474 against 0.838; *p ≤* 0.003 in every decomposition–model combination). Internally, the same synthetic spectra looked considerably more successful: they were classified correctly by real-trained models (Real2Syn, mean accuracy 0.874), and adding them to the training set gave a high cross-validated accuracy (Mixed2Real, 0.903). Only the external tier, and the Syn2Real strategy (mean 0.553, itself an optimistic figure; see Limitations), exposed that the synthetic data carried little information beyond what the real data already contained.

### 4.2 Synthetic Spectra Are Classifiable but Not Substitutable

The asymmetry between Real2Syn (0.874) and Syn2Real (0.553; 0.500 when features are selected on the synthetic training set rather than on the real test spectra) was present in 45 of the 48 settings and was not confined to a particular task, echo time, generative model or decomposition. A real-trained classifier can label synthetic spectra correctly because those spectra fall inside the decision regions it learned from the real data. The converse fails because the synthetic spectra do not span the real distribution: a classifier trained on them learns boundaries that are placed too tightly around the synthetic class centres and misclassifies the more variable real spectra. This is the pattern expected from a generator that is accurate about where each class is centred but too narrow about how far real spectra stray from that centre. It is not the pattern expected from an inaccurate generator, which would degrade Real2Syn as well.

Three observations from the generative pipeline support this interpretation. First, the selection criterion favoured very simple generators: a single-component GMM (one Gaussian per class) was chosen in 7 of the 12 task-specific configurations, and small KDE bandwidths were chosen throughout (Section 3.1). Second, at the selected GMM setting the synthetic spectra lay at least as close to the real class means as the cNMF reconstruction of the real weights did, in every configuration, and at only about half to two-thirds of the distance at LTE for five of the six tasks (e.g. MM vs ME, LTE: 3.6*×*10*^−^*^4^ against 7.8*×*10*^−^*^4^). This comparison also shows that the real-weight reconstruction is not a floor for the MSE: a generator can score better than the real data it was fitted to precisely by being less variable than it. A third observation concerns the sampler itself: more than half of the sampled weight vectors were rejected for having a non-positive component (Table 5), so the synthetic weights come from a truncated version of the fitted distribution, which further concentrates them. Synthetic spectra that are closer to the class mean than real spectra are less variable than the data they are meant to represent. This is a methodological point worth foregrounding: fidelity to the class mean and usefulness as training data are not the same objective, and a criterion that optimises the former can actively select against the latter.

### 4.3 Why Augmentation Did Not Improve External Generalisation

Several explanations are consistent with our results, and they are not mutually exclusive; the present experimental design cannot separate them.

#### Synthetic data add volume, not diversity

If the generated spectra lie within the envelope of the real training spectra, as the domain-gap findings suggest, then adding them to a linear classifier trained on ten selected features changes little, which is what we observed: Real+Syn2Ext left accuracy unchanged in 4–5 of the 12 settings and moved it in either direction in the rest.

#### The baseline was already strong

The Real2Ext baseline reached a mean external accuracy of 0.838, and 1.000 for A2 vs NO at LTE with the split decomposition, leaving limited headroom in the easier tasks. Improvement was more plausible in the harder three-class tasks, but the effects there were of both signs and comparable in magnitude.

#### Limited statistical power

With twelve paired settings per comparison, the 95% confidence intervals on the mean difference had half-widths of roughly 0.03–0.04 and upper limits of at most +0.053. Improvements larger than about five percentage points can therefore be regarded as unlikely, but smaller benefits could not have been detected.

### 4.4 Task-Specific versus Joint Decomposition

No comparison between the joint five-class decomposition and the task-specific decompositions was significant (all *p ≥* 0.147; Table 10). A practically useful reading is that one shared decomposition can serve several diagnostic tasks without a measurable cost in average performance, which simplifies deployment. The setting-level differences were, however, sometimes large and of sign, up to +0.42 and *−*0.54 for Syn2Ext, and even the Real2Ext baseline differed by up to about 0.1 owing only to which cases survived outlier removal. We interpret this as evidence of instability in individual configurations rather than of a real advantage in either direction.

### 4.5 Comparison with Prior Work

In a recent review by the MRS Synthetic Data Working Group [31], only a small number of studies to date have created synthetic magnetic resonance spectroscopic data specifically tailored to clinical scenarios such as brain tumour classification. The closest MRS-specific precedent for generating spectra is the comparison of linear, non-linear and deep generative models by Olliverre *et al.* [6]. Their evaluation used a held-out subset of the same dataset, whereas ours uses an independently acquired multi-centre cohort; the weaker benefit we report may therefore reflect the harder test rather than a weaker generator, and our own internal results (Mixed2Real, 0.903) would have supported a more favourable conclusion. cNMF has been used for source extraction and decomposition-driven classification of MRS data [13–15]; we extend the same representation to a generative role and, to our knowledge, provide the first evaluation of that use against an independent external cohort. Our real-data baseline can be situated against the earlier INTERPRET-to-eTumour evaluation [5].

## 5 Conclusions

This study evaluated whether synthetic MRS spectra, generated by cNMF decomposition and resampling of the resulting weight distributions, can improve the external generalisation of brain tumour classifiers. The synthetic spectra produced by this approach are internally coherent and are readily classified by models trained on real data, confirming that the generative procedure captures genuine class structure. They did not, however, improve classification performance on an independently acquired multi-centre cohort relative to training on real data alone, and they proved a poor substitute for real training data when used in isolation. The pattern of results points to a generative distribution that is faithful to each class’s central tendency but too narrow to supply the variability that augmentation requires, a shortcoming we interpret as arising from the criterion by which the generative models were selected rather than from the decomposition itself, although the present design does not test this directly.

Two contributions follow. The first is methodological: to our knowledge, this is the first evaluation of cNMF-based weight-space resampling as a generative strategy for MRS augmentation, extending a decomposition previously used for source extraction and classification into a generative role, and identifying the mismatch between reconstruction fidelity and augmentation utility as the critical design decision in doing so. The second is evaluative, and applies beyond the particular method studied here. Had this work relied on internal cross-validation alone, the same synthetic data would have appeared to succeed (although the internal estimates are themselves optimistic); it was only under independent external validation that its limitations became apparent. We therefore argue that independent, ideally multi-centre, external cohorts should be treated as a requirement rather than a refinement when assessing synthetic data augmentation in MRS, and we suggest that generative models intended for augmentation be selected on their distributional coverage rather than on their fidelity to a class average.

## Data Availability

All data produced are available online at https://doi.org/10.34810/DATA3818

## Acknowledgements

PID2024-155606OB-I00, Agencia Española de Investigación (AEI), Spain. 2025-STEP-00154, AGAUR, Generalitat de Catalunya. FP6-2002-LIFESCIHEALTH-503094, European Union. FP5-IST-1999-10310, European Union. MEC SAF2005-03650 Ministerio de Educación y Ciencia, Spain. Centro de Investigación Biomédica en Red en Bioingeniería, Biomateriales y Nanomedicina (CIBER-BBN [http://www.ciber-bbn.es/en, accessed on 1 November 2026], CB06/01/0010), an initiative of the Instituto de Salud Carlos III (Spain) co-funded by EU Fondo Europeo de Desarrollo Regional (FEDER). Spanish AEI PID2019-104551RB-I00 grant. Xartecsalut, 2021 XARDI 00021

## Declarations

### Ethics statement

Data were acquired in the context of FP5-IST-1999-10310, FP6-2002-LIFESCIHEALTH-503094 and MEC SAF2005-03650, according to medical ethics regulations, with the Helsinki Declaration and the Spanish “Ley Orgánica de Protección de Datos de Caŕacter Personal (LOPD), Ley Orgánica 15/1999” and the “95/46/EU directive on data protection, December 13th, 1999”. All patients or their legal representatives signed informed consent forms, agreeing to the study and to the use of their deidentified (anonymized) data for research.

### Author contributions

Conceptualization, M.J.-S., A.V.; methodology, M.M., A.V. and M.J.-S.; software, M.M. and L.T.; data curation, M.J.-S. and M.M.; writing—original draft preparation, M.M, A.V. and M.J.-S.; writing—review and editing, all co-authors; funding acquisition, M.J.-S., A.V. All authors have read and agreed to the published version of the manuscript.

